# EARLY-PREG: A preconception longitudinal cohort of women seeking pregnancy to investigate maternal-embryonic molecular signalling during the peri-implantation window

**DOI:** 10.1101/2025.05.12.25326372

**Authors:** Elard S. Koch, Denisse Avila, Guillermo Nourdin, Veronica Latapiat, Barbara Antilef, Estefanía Contreras, Mauricio Hernandez, Juan F. Stecher, Cristian Vargas

## Abstract

**BACKGROUND:** Maternal-embryonic signalling involves intense and complex molecular exchange between the maternal reproductive system and the early embryo. This interaction begins after fertilisation but is poorly understood before implantation. EARLY-PREG is structured as a bidirectional prospective–retrospective preconception open cohort designed to characterise time-dependent molecular signatures of maternal–embryonic communication during the earliest stage of pregnancy. The cohort supports a dynamic and structured longitudinal biobank of maternal biological fluids, cells and tissues-derived samples collected during the peri-implantation window.

**STUDY DESIGN:** Participants were recruited in three rolling waves from 2017 to 2024 so far, in Concepcion, Chile. Healthy women trying to conceive were enrolled and completed a survey with health, lifestyle, and sociodemographic data. Menstrual cycles were prospectively monitored with ovulation and fertile window ascertainment (ultrasound, fertility monitor, and/or LH strips) and followed for a maximum of six consecutive cycles or until implantation of a viable embryo exposure occurred.

Each cycle was synchronised within a longitudinal repeated-measures design incorporating systematic day-by-day sampling anchored to ovulation (day 0). Biological samples include cervicovaginal fluid (CVF), urine, saliva, and blood. In addition, cervical brushings were collected during the peri-implantation window (days 12–14 post-ovulation) and on day 21 post-ovulation. Retrospective temporal alignment of ovulation and defined physiological windows was performed using hormonal curves (LH, oestradiol, progesterone and beta-hCG) measured in stored blood and urine samples.

**RESULTS TO DATE:** At present, 1,183 women have been contacted, of whom 223 met all eligibility criteria. Among participants trying to conceive, 129 completed at least one full longitudinal repeated-measures cycle; 35 achieved full-term pregnancies and 17 experienced early pregnancy loss (ELP). In addition, 40 abstinent and 5 sterilised women completed the protocol.

To date, 292 menstrual cycles have been fully documented and sampled. 52 cycles correspond to conception cycles and 240 cycles to the absence of embryo implantation. Among the latter, 31 cycles were classified as non-conception counterfactual cycles and 209 to non-counterfactual cycles. The biorepository encompasses maternal biological samples collected during the first 2 weeks after ovulation across all documented cycles. Biospecimen collection compliance exceeded predefined protocol thresholds for most sample types.

**CURRENT AND FUTURE DIRECTIONS:** The cohort currently supports longitudinal proteomic analyses within a within-individual counterfactual framework, comparing the exposure to a viable embryo implantation (conception cycle) with the absence of embryo implantation (non-conception cycle) to characterise time-dependent molecular signatures of early pregnancy. Additional outcomes of the cohort include implantation failure and early pregnancy loss. A fourth rolling recruitment wave is planned for 2026 to characterise time-dependent immunophenotypic variation in maternal peripheral blood mononuclear cells (PBMCs) during early pregnancy stages.

**STUDY FUNDING:** This work forms part of the EARLY-PREG preconception open cohort and has been supported by research grants awarded by Fundación de Investigación San Ramón (FISAR), Chile. The pilot study and first recruitment wave were supported by grants #MEL109112011 and #MEL109112011R4 awarded to E.S.K., C.V., and J.F.S. The second recruitment wave was supported by grants #MEL109112011R5 and #MEL131032017R1 awarded to E.S.K. The third wave was supported by grant #MEL205062018 awarded to E.S.K. and M.H. Current funding for the fourth recruitment wave and mass spectrometry research on maternal CVF is supported by grant REH042024-01 awarded to M.H., G.N., and E.S.K.

**TRIAL REGISTRATION NUMBER:** NCT07358026.

## 1. Introduction

Maternal-embryonic signalling is critical for the establishment of a successful pregnancy. The period after fertilisation and before implantation, known as the *pre-implantation* period, was previously thought to be silent. However, evidence suggests that this communication lays the groundwork for pregnancy (Barnea *et al*., 2012). During this period, intense and complex molecular exchange occurs between the early embryo and the maternal endometrium (Lane *et al*., 2014). At present, researchers have not fully characterised this interaction (Lynch *et al*., 2006; Benagiano *et al*., 2023a, 2023b). This maternal–embryonic dialogue involves a complex interaction between the embryo and the mother, starting at fertilisation, termed the ultra-early stages of pregnancy (Hill, 2001; Barnea, 2004; Singh *et al*., 2011). Through this communication, the embryo can modulate the maternal response during key phases, such as apposition, adhesion, invasion, and decidualisation in the endometrium, through extensive exchange of signals (Somerset *et al*., 2004; Barnea, 2007; Lédée *et al*., 2007; Fazeli, 2011). The success of each of these events is essential for advancing to the next stage. However, the regulatory mechanisms that govern this maternal–embryonic interaction are still not completely understood (Cha *et al*., 2012).

During the pre-implantation period, the uterine fluid potentially facilitates the transfer of vital information between mother and the developing embryo (Zhang *et al*., 2017). In this context, the presence of molecules involved in the interaction between the embryo and the maternal environment, such as the early pregnancy factor (EPF, Morton *et al*., 1982), preimplantation factor (PIF) (Barnea *et al*., 1994) and PAF (embryo-derived platelet-activating factor), has been reported (O’Neill, 1992, 2005). EPF and PIF are involved in the maternal immune response, which is crucial for the viability of early embryos and implantation (Nahhas and Barnea, 1990; Morton, 1998; Barnea *et al*., 2012, 2014; Santos *et al*., 2021). It has been postulated that EPF originates from the mother during the preimplantation period and shifts to embryonic origin after implantation (Nahhas and Barnea, 1990; Morton, 1998; Barnea *et al*., 2012, 2014; Santos *et al*., 2021). PIF secreted by the embryo has recently been suggested to enhance the decidualisation process and the production of endometrial factors that limit trophoblastic invasion (Santos *et al*., 2021). Moreover, a third factor known as PAF has been described; its activity as an embryotrophin would mediate the transport of the embryo to the uterus (O’Neill, 1992, 2005). In addition, recent studies have shed light on new forms of embryo–maternal communication via the delivery and/or exchange of extracellular vesicles (EVs) and mobile RNAs. Furthermore, EVs play a role in generating an immunosuppressed environment during embryo‒mother interactions (Simon *et al*., 2018; Kaminski *et al*., 2019; Das and Kale, 2020), allowing the embryo to counteract the maternal immune response (Burnett and Nowak, 2016). Nevertheless, further investigation is needed to fully understand the maternal–embryonic dialogue.

Embryo‒mother communication in humans is a unique and challenging-to-model process. Commonly, *in vivo*, *ex vivo*, and *in vitro* models have been developed to simulate certain phases of the implantation process and its related events (Dimova *et al*., 2024). Despite advances in experimental strategies, understanding the mechanisms of embryo‒maternal crosstalk faces inherent limitations that affect how well its findings can be extrapolated to humans. Notably, variation between humans and animal models has been observed in decidualisation, the timing and type of implantation, attachment, trophoblast subpopulations, and the depth of extravillous trophoblast invasion (Muter *et al*., 2023; Dimova *et al*., 2024). Moreover, *in vitro* models designed to replicate the implantation process have been developed using immortalised cell lines or cancer cells, which may not accurately represent normal physiological conditions (Shibata *et al*., 2024). Thus, the maternal–embryonic dialogue during early stages requires further research using novel strategies tailored to human physiology.

In recent years, proteomic profiling in longitudinal cohort studies has emerged as a modern strategy for understanding complex biological processes (Romero *et al*., 2017; Aghaeepour *et al*., 2018; Yohannes *et al*., 2022). These advancements have been driven by innovations in Mass Spectrometry (MS), which have allowed the integration of proteomic data with pathway analysis. Proteomics has proven to be a reliable and highly sensitive method for biological research (Cox and Mann, 2011; Geyer *et al*., 2017), particularly because of its ability to analyse biological fluids (Bader *et al*., 2023). Proteomic profiling, combined with longitudinal sampling of biological fluids, is a non-invasive alternative to monitoring processes, such as the embryo‒mother molecular exchange of signals.

Longitudinal proteomic profiles during pregnancy in healthy women have been investigated in a few cohort studies, starting from the 8th week of gestation. Although these studies have performed proteomic analyses during pregnancy, to date, no studies have characterised the first two weeks of gestation from the time of conception (Romero *et al*., 2017; Hedman *et al*., 2020). Consequently, the earliest stages of maternal–embryonic interaction remain poorly understood, particularly without altering, manipulating, or intervening in their natural environment.

To address this gap, we designed EARLY-PREG, a preconception, longitudinal, bidirectional, and counterfactual cohort study. EARLY-PREG investigates maternal–embryonic communication during the first two weeks after conception through a biorepository of maternal biological fluids and tissues collected frequently throughout the preimplantation period and into pregnancy. Crucially, the value of this cohort depends on the implementation of advanced analytical strategies that can sensitively and reproducibly capture the molecular complexity of cervicovaginal fluid (CVF), a biofluid central to reproductive physiology (Zegels *et al.,* 2010; *Fernandez-Hermida* et al., 2018). In a companion study, we validated a data-independent acquisition parallel accumulation–serial fragmentation (dia-PASEF) method for CVF large scale longitudinal proteomics with repeated measures over time identifying specific physiological windows, demonstrating superior reproducibility, reduced missing values, and improved quantification of low-abundance proteins compared with traditional approaches (Hernandez *et al.,* 2025). Incorporating this methodology into EARLY-PREG ensures that the unique temporal resolution of the cohort can be fully leveraged to discover dynamic proteomic signatures of early pregnancy.

This protocol article completely outlines the study design and provides an overview of the data collected so far, including baseline participant information, reproductive outcomes, menstrual cycle characteristics, biological samples, and protocol performance.

## 2. Cohort description

### 2.1 Study design

The preconception study employs an open longitudinal, bidirectional, and counterfactual cohort design, which is prospective when successive day-by-day samples are collected and retrospective for analysis once pregnancy is confirmed. In addition, this design allows each participant to serve as their own control (counterfactual), enabling the comparison of non-conception and conception cycles while minimising interindividual variability, as detailed in Figure 1.

**Figure 1.**
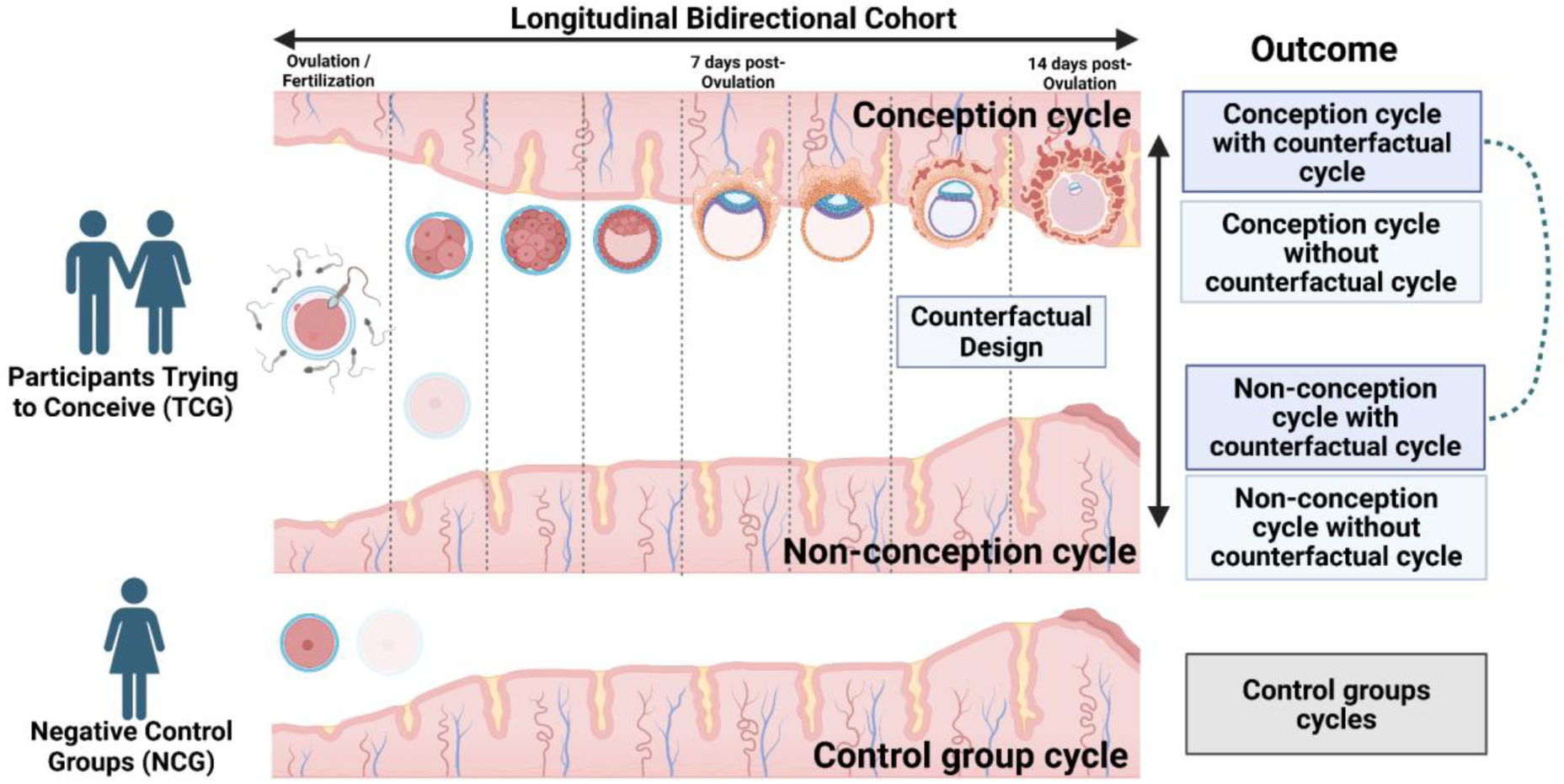
EARLY-PREG cohort design. The figure represents our two main study groups: Participants trying to conceive (TCG) and a negative control group (NCG), and illustrates the endometrium in three distinct scenarios — conception cycle when pregnancy occurs, non-conception cycle when pregnancy does not occur, and control cycle without pregnancy due to abstinence and/or sterilisation— along with the anticipated outcomes.

### 2.2 Bidirectional longitudinal design

Given the study’s objective, a longitudinal prospective follow-up with repeated measures was established, beginning before the onset of pregnancy in the study participants. Measurements were planned to continue while couples sought pregnancy throughout the menstrual cycles, during pregnancy, and until delivery. A bidirectional or two-way component was incorporated into longitudinal follow-up for sampling. Consequently, the occurrence of pregnancy serves as a milestone for the retrospective hormonal analysis (LH, oestradiol, progesterone and beta-hCG) of the stored samples to correct and synchronise specific time periods of interest, such as the fertile window, the preimplantation window, and the implantation window.

### 2.3 Counterfactual approach

Counterfactual models use hypothetical scenarios to estimate the outcomes that individuals would have experienced if they had been exposed to a different intervention than the one they received. The counterfactual design of EARLY-PREG enables a comparison of the systemic and localised effects of embryo appearance and implantation in an individual (the factual) against the physiological state of the same individual without experiencing pregnancy (the counterfactual). Comparison of an early-in-life event with a late-in-life event was restricted to a range of up to six menstrual cycles from the start of active pregnancy seeking (Eichler *et al*., 2016).

### 2.4 Setting and recruitment

The EARLY-PREG cohort consists of healthy couples trying to conceive and women not trying to conceive. Since 2017, a total of 1,183 women have been invited to participate through word of mouth, invitations from gynaecologists, and midwives in private practices, public hospitals, and Family Health Centres (CESFAMs) in the Biobio region of Chile.

Women interested in participating in the study went through a pre-selection phase to receive general information about the study in a consultation room at the *Sanatorio Aleman* Clinic in Concepcion, Chile. Potential participants were subsequently interviewed by the midwife to address any questions from the recruitment team and to request participation in the study from the women and their partners (in the case of couples trying to conceive), which was formalised through corresponding informed consent.

### 2.5 Selection Criteria

Eligibility requirements common to all women include being between 18 and 40 years of age, not being pregnant, normal colposcopy, body mass index (BMI) between 18 and 29, having regular menstrual cycles (21–35 days), and absence of chronic diseases (hypertension, diabetes mellitus, cancer, depression, personality disorder, thyroid pathology, polycystic ovary syndrome, or hyperprolactinemia). Pregnant women and those with a history of alcoholism, infertility treatment, endometriosis, pelvic inflammatory disease or pelvic surgery, or allergy to latex or silicone were excluded. In addition, eligibility criteria were established for the partners of women trying to conceive. Women were included if their partners were males between 18 and 40 years old, without pathologies (diabetes mellitus, depression, personality disorder, or cancer), were not taking chronic medication, without erectile disorders, BMI between 18 and 29.9, moderate alcohol use, and with no recreational drug use. The exclusion criteria were working in contact with pesticides, a history of erectile dysfunction, mumps in adulthood, chronic diseases, and psychological problems.

### 2.6 Study groups

Figure 2 shows the recruitment flowchart for the entire EARLY-PREG cohort. The women enrolled so far in the study are divided into two main categories: women trying to conceive (TCG) and women who are not seeking pregnancy, which represents a negative control group (NCG). The negative control group is further categorised into two subgroups: women practising abstinence and women who were sterilised. Within the sterilised women, there are additional subdivisions: women with sexual abstinence and women without sexual abstinence. These negative control groups will be used to establish proteome libraries under initial conditions that differ from those of conception or the ultra-early pregnancy stage.

**Figure 2.**
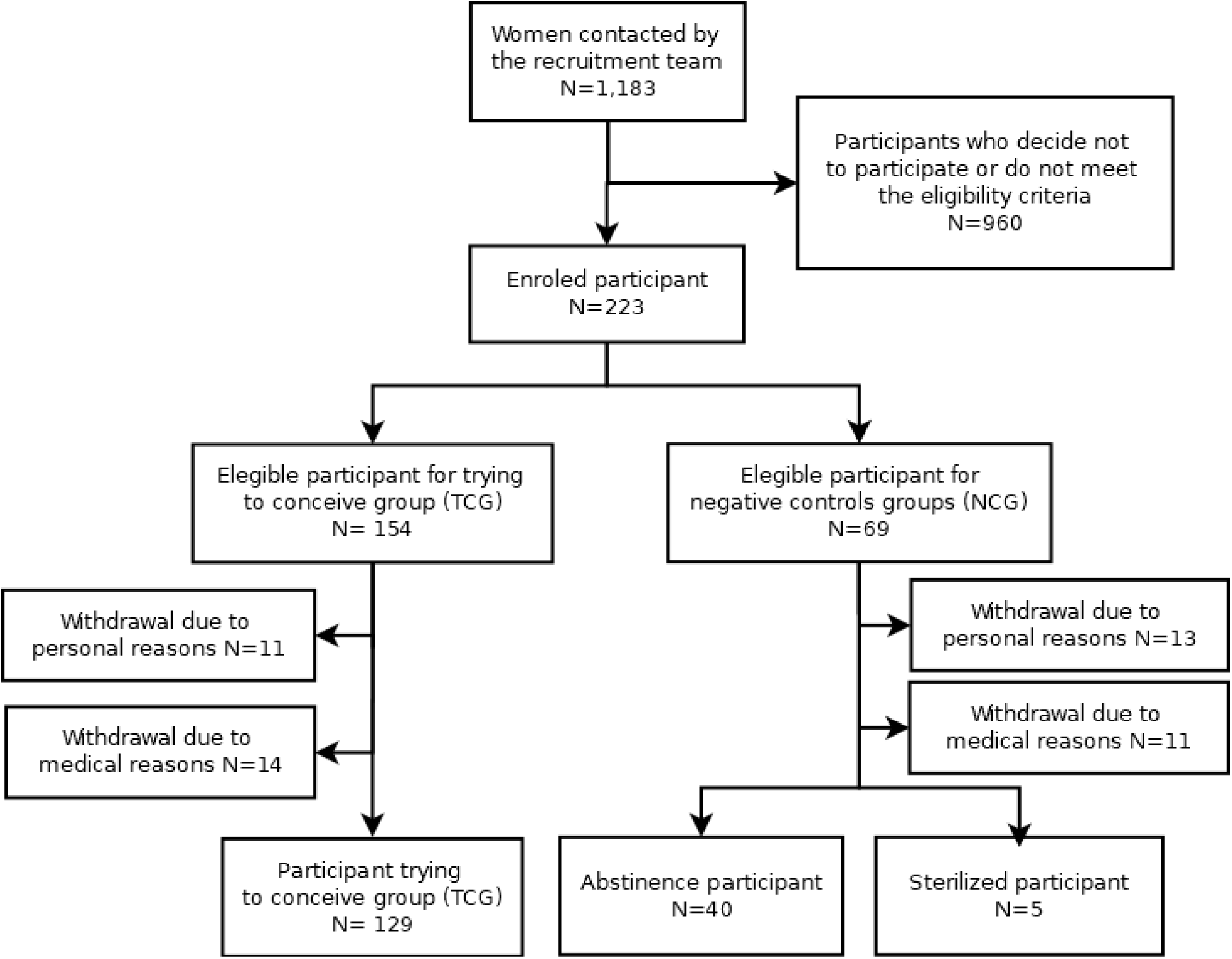
Flow chart of the EARLY-PREG open cohort design.

Throughout the study, participants are monitored during their cycles through the collection of various biological samples, including blood, urine, saliva, CVF, and cervical brushing, for both groups. The NCG is followed for a single complete cycle, whereas the TCG is monitored over multiple cycles (up to six cycles), with serial collection of biological samples. If a pregnancy is identified in the TCG study group, monitoring continues until the pregnancy concludes.

A total of 1,183 women have been contacted so far and were recruited during the screening phase. A total of 223 participants were selected and agreed to participate in the study through consent forms and enrolment in the protocol. During the study, 49 participants withdrew from the protocol (21.97%) (Figure 2). The main reasons for withdrawal were classified as personal or medical. A total of 292 menstrual cycles from 174 women have been monitored so far, with 35 achieving full-term pregnancies.

### 2.7 Patient and public involvement

The study participants were not involved in the design, conduct, reporting, or dissemination of our research. They were not directly involved in the study’s design, the development of the research questions and outcome measures, or the recruitment and execution.

The protocol was reviewed and approved by the ethics committee of the Servicio de Salud Concepcion, Biobio region, Chile (CODE: 17-03-06). The committee’s role was to advise on the ethical and operational aspects of the study.

### 2.8 Questionnaire survey

At enrolment, both female and male participants complete a baseline questionnaire survey, which is conducted and recorded by a midwife from the research team. The questionnaire includes sections for females, males, and sociodemographic information. Both women and their male partners (when applicable) are asked about their health (e.g., BMI, allergies, chronic diseases, and use of chronic medications) and lifestyle (e.g., smoking habits, alcohol consumption, drug use, habit of coffee drinking, and physical activities). Additionally, women are asked about contraceptive use, reproductive and general obstetric history, and dietary habits. The sociodemographic section includes questions on marital status, age, educational level, occupation, and income.

## 3. Methods

### 3.1 Main Outcomes

The three main outcomes in the cohort are defined according to menstrual cycles in which conception is achieved and those in which conception is not achieved. Below are their respective clinical definitions:

- **Number of menstrual cycles leading to full-term pregnancies:** This outcome refers to the menstrual cycle in which the ovum is fertilised, leading to pregnancy with a full term live birth. The conception cycle requires beta-hCG levels above the clinical threshold for a positive pregnancy test, which is determined in peripheral venous blood on the 14th day post-ovulation.
- **Number of menstrual cycles not leading to pregnancy:** This outcome refers to a menstrual cycle in which pregnancy does not occur. When referring to the same individual, it is, by definition, considered the counterfactual to the conception cycle described above. A non-conception cycle is confirmed by a clinically negative beta-hCG test, which is determined in peripheral venous blood on the 14th day post-ovulation. This cycle is characterised by the absence of a clinical pregnancy.
- **Number of menstrual cycles leading to early pregnancy loss:** This outcome refers to the menstrual cycle in which the ovum is fertilised, leading to a miscarriage until 12 6/7 weeks. The conception cycle requires beta-hCG levels above the clinical threshold for a positive pregnancy test, which is determined in peripheral venous blood on the 14th day post-ovulation.

### 3.2. Recruitment, biospecimen collection and follow-up

#### 3.2.1 Recruitment phase

Approval by the ethics committee of the Servicio de Salud de Concepcion was obtained before the recruitment stage. Participants were recruited through medical consultations, health centres, and referrals from doctors and midwives. An initial interview was conducted to explain the details of the study and assess eligibility based on the inclusion and exclusion criteria. The study was discussed in a private room at the respective health centres, and informed consent was obtained.

Each participant undergoes a clinical evaluation, including a physical examination and speculoscopy. A Papanicolaou (Pap) test was administered to women who had not recently undergone one to screen for cervical cancer. Instructions for cycle follow-up were provided both orally and in writing.

#### 3.2.2 Data collection

Personal, clinical, and follow-up data have been collected in individual files. Each participant is anonymised and assigned a unique code to ensure data confidentiality.

Additionally, the midwives responsible for recruitment and follow-up transfer the information from the physical records to a digital document on password-protected computers belonging to MELISA Institute.

The collected samples are registered in a digital inventory stored on password-protected computers belonging to MELISA Institute. Each sample is registered using the unique code assigned to each participant to ensure confidentiality.

#### 3.2.3 Ovulation and fertile window ascertainment

Although conception cannot be directly observed, estimating the day of ovulation allows researchers to narrow the time frame during which fertilisation is most likely to occur (Wilcox *et al*., 1995). Furthermore, the estimation of ovulation day allows the targeted collection of biological samples to characterise and investigate the fertile window in non-conception cycles and the periimplantation window in conception cycles. The EARLY-PREG cohort has undergone three recruitment waves so far, each defined by variations in the clinical method used to estimate the day of ovulation and fertile window, following the proposed schema in Figure 3A. The pilot study and first wave relied on ultrasound to determine ovulation. The second wave employed a portable fertility monitor (Clearblue Digital Ovulation test). In the third wave, commercial LH strips were used to infer ovulation from urine samples. All recruitment waves included a professional instructor who taught women how to estimate their fertile window with the Billings method. Detailed, hand-filled fertility charts were used for each cycle. A nurse or midwife in daily contact with each couple provided reproductive counselling.

**Figure 3.**
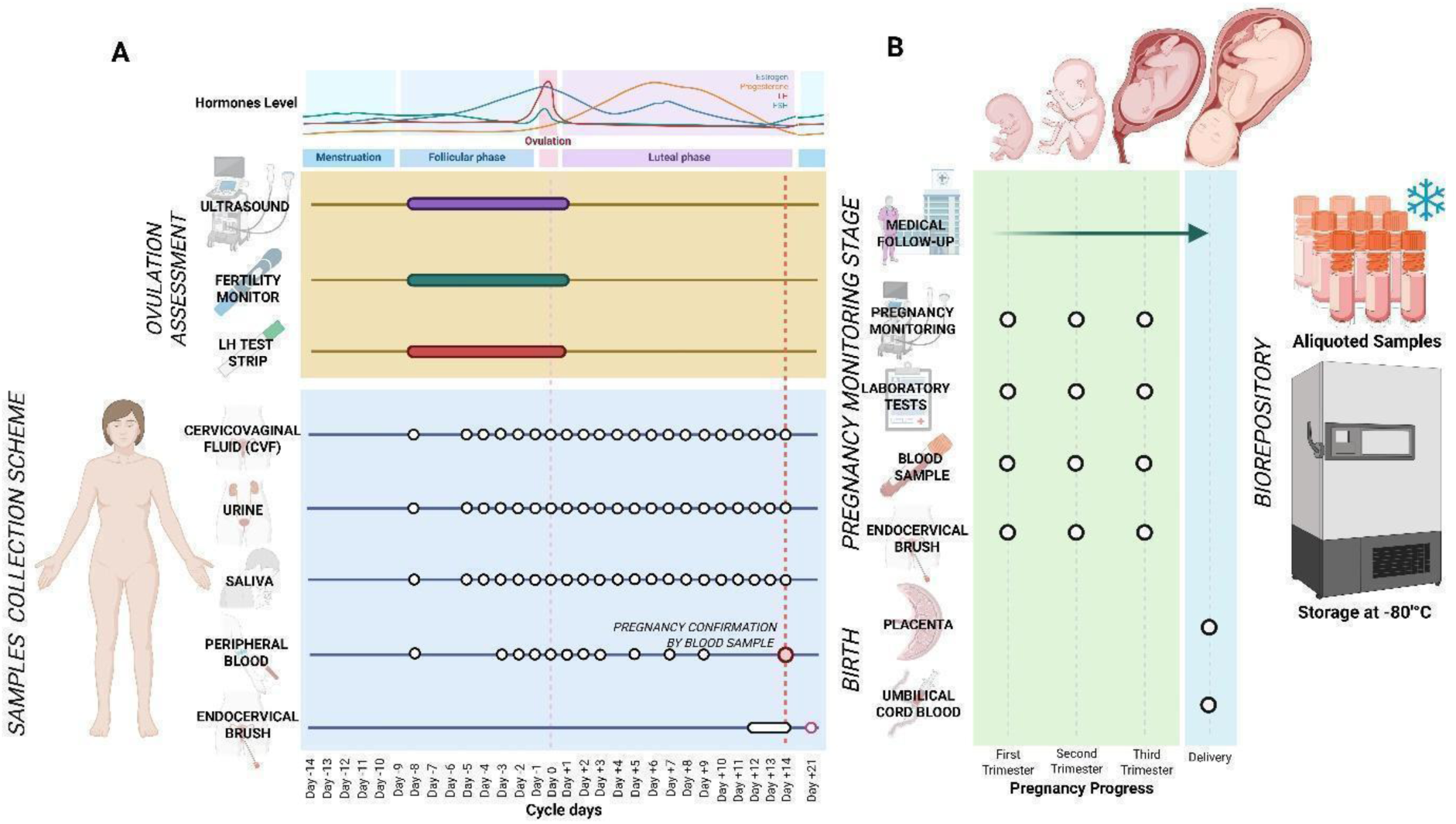
EARLY-PREG cohort workflow scheme. A. Systematic collection of biological samples (CVF, saliva, blood, cervical brushing, and urine) from participants actively trying to conceive and participants not trying to conceive. In addition, cervical brushing is performed on day 21 post-ovulation for each participant with a clinically confirmed pregnancy. Cycle days are expressed in distance from ovulation day (day 0). B. For participants who achieve pregnancy, additional samples, such as umbilical cord blood and placenta, are obtained at delivery.

#### 3.2.4 Correction and synchronisation of counterfactual cycles

Given the high variability of menstrual cycles, identifying the day of ovulation is a key issue in the EARLY-PREG cohort. In fact, this day, estimated with a precision margin of approximately 12 hours, is considered the most reliable proxy for the day of conception, or day 0, in the cohort of pregnant women. The two-way or bidirectional design used in the present preconception cohort offers a critical temporal framework for studying ultra-early pregnancy events with improved accuracy for multiple non-conception and conception menstrual cycles in a single individual following a counterfactual model. The fertile window typically includes the five days preceding ovulation, the day of ovulation itself, and the day following the estimated ovulation (Wilcox *et al*., 1995). While all the above methods offer a pragmatic approach to clinically estimating the fertile window and providing reproductive counselling at the specific period of the menstrual cycle during which conception is possible, these methods, however, are inaccurate and do not allow synchronisation of multiple cycles for research purposes.Therefore, to address this major issue, comprehensive laboratory analyses of hormone curves—including LH and beta-hCG in urine, and oestradiol and progesterone in serum—are retrospectively conducted on stored samples to more accurately synchronise the day of ovulation across multiple counterfactual cycles and/or to identify the preimplantation window within a conception cycle more precisely. An example of a multi-cycle correction method used to synchronise ovulation (LH-based approach) and implantation (beta-hCG-based approach) used in the cohort is presented in Figure 4. The combination of these methods allows for a more precise isolation of specific time windows of interest in counterfactual cycles within the same individual.

**Figure 4.**
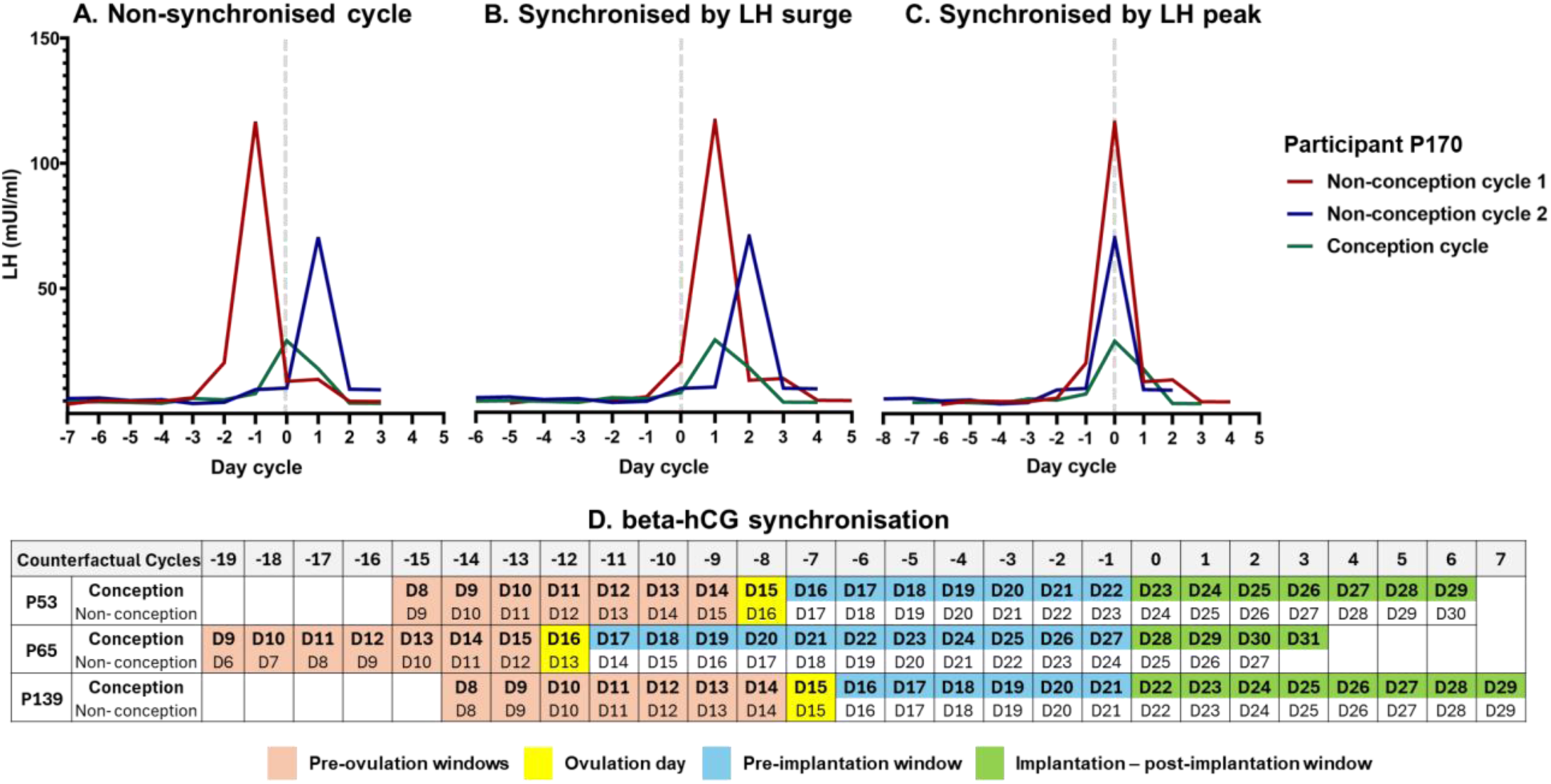
Synchronisation methods based on ovulation and the implantation process using urine samples. The figure illustrates two types of synchronisations: one based on luteinising hormone (LH) concentration for ovulation assessment, and the other on beta-hCG concentration for embryo implantation. The red, blue, and green lines represent three LH concentration profiles from cycles of the same participant. Panels A, B and C show synchronisation based on ovulation day and LH concentration for two consecutive non-conception cycles and a third conception cycle in the participant P170 of the cohort. In panel A, the cycles are non-synchronised. In panel B, the cycles are synchronised using the LH surge strategy proposed by Godbert et al. (2015). In panel C, the cycles were synchronised through the LH peak concentration. Panel D presents synchronisation based on beta-hCG presence in counterfactual cycles of three participants, P53, P65, and P139 of the EARLY-PREG open cohort. The cells represent a specific day (D) of the menstrual cycle for each participant, aligned according to the beta-hCG synchronisation approach. The colours in the table indicate different phases of the cycle: pre-ovulation window (orange), ovulation day (yellow), pre-implantation window (blue) and implantation/post-implantation window (green). This alignment enables a standardised comparison of parallel time windows across counterfactual cycles in the same individual.

#### 3.2.5 Biological material handling and storage

The strategy for collecting biological samples used in the EARLY-PREG cohort is intensive and challenging. The protocol is based on daily serial collection of CVF, urine, saliva and blood. A cervical brushing sample is collected between days 12 and 14 post-ovulation in all participants. Additionally, blood samples and cervical brushings are collected from pregnant women on day 21 after ovulation and at each trimester of pregnancy. Finally, placenta and umbilical blood are obtained at childbirth (Figure 3B). A longitudinal repeated-measures design has been performed during the phases of interest, including the follicular, periovulatory, and luteal phases. The specific details about the collection protocols for each sample are described below:

##### Blood

Four tubes of venous blood are drawn using BD Vacutainer tubes: two with EDTA (ethylenediaminetetraacetic acid) for proteomic analysis, peripheral blood mononuclear cell extraction, and platelet count, and two with a separator gel and coagulation activator for beta-hCG and hormone analysis. The samples are collected by nursing staff and transported to the laboratory in a cooler box with cold gel packs. The plasma is obtained by centrifugation at 2000 × g for 15 minutes and stored at −80 °C.

##### Urine

The participants are trained to collect morning urine at home in a sterile 60 mL container, discarding the initial stream. Instructions are given to keep the samples refrigerated (∼4 °C) until daily collection. The urine samples were transported to the laboratory in a cooler box with cold gel packs. In the laboratory, the samples were centrifuged (600 × g, 10 minutes), and the supernatants were stored at −80 °C for hormone analysis (oestradiol, FSH, LH, progesterone, and beta-hCG).

##### Saliva

The participants collect ∼1 mL of saliva at home using Salimetrics SalivaBio passive collectors before eating and after brushing their teeth in the mornings. Instructions from the manufacturer are provided to participants for the collection of samples. Saliva samples are collected daily and transported to the laboratory in a cooler box with cold gel packs. The samples are stored directly at −80 °C in cryotubes.

##### Cervicovaginal Fluid (CVF)

The participants collected CVF using a silicone menstrual cup worn for two hours during the first hours of the morning. Cups are washed with a pipette using a buffer that contains PBS, physiological serum, and protease inhibitors and then transferred into a borosilicate tube. The samples are retrieved daily and transported to the laboratory in a cooler box with cold gel packs. These samples are subsequently processed (centrifuged and aliquoted) and then stored at −80 °C.

##### Cervical Brushing

Cervical brushing samples are collected from the cervical canal area by trained staff via a Rovers Cervex-Brush. The brush is then placed in a 50 mL tube containing medium enriched for trophoblastic cells (RPMI) with foetal bovine serum (FBS) and antibiotics. The samples are transported to the laboratory within 4 hours of collection via a cooler box with cold gel packs. Brushes are washed with PBS and then centrifuged. The collected pellet is resuspended in 400 μL of PBS and transferred to a microcentrifuge tube. Subsequently, 100 μL of 4% paraformaldehyde is added, and the sample is finally stored at 4 °C for further analysis.

##### Umbilical cord blood

At birth, the obstetrician collected blood from the cord, umbilical vein, and umbilical artery using BD Vacutainer tubes with EDTA. The sample collection times are recorded, and the samples are then immediately sent to the laboratory. The umbilical cord blood is transported at room temperature (20 °C–25 °C) in a transport box. Sample processing involved centrifugation to separate peripheral blood mononuclear cells using Ficoll gradients. The cells are stored in RPMI medium supplemented with 20% FBS and 5% DMSO. Initially, the samples were stored for two weeks at −80 °C and then transferred to liquid nitrogen for long-term storage.

##### Placenta

Obstetricians collected placenta samples at the time of birth. The tissue was sectioned under sterile conditions, placed in a sterile container, and then transported to the laboratory in a cooler box with cold gel packs. The samples were preserved in RPMI medium supplemented with 20% FBS and stored at −80 °C for further analysis.

#### 3.2.5 Follow-up stage

The length of follow-up is variable between individuals and study groups. The TCG is monitored for up to six menstrual cycles or until pregnancy, whichever comes first. In the case of pregnancy, follow-up is extended to the end of the pregnancy. The NCG is followed for a single menstrual cycle.

For all the participants, the follow-up phase begins on the first day of menstrual bleeding after enrolment, which is reported by the participant to the team via a phone call. This day marks the establishment of the cycle start date and serves as a milestone for scheduling visits and daily phone calls with participants. The first visit takes place between the third and fifth days of the cycle, when trained staff collect urine and peripheral blood samples to exclude pregnancy through a beta-hCG test. An ultrasound is performed during this visit for gynaecological evaluation of uterine and ovarian structures (this assessment is performed once). Additionally, the clinical staff provides counselling to couples regarding sexual intercourse during the fertile window of each menstrual cycle.

For monitoring, days 6 and 9 of the cycle include visits for collecting peripheral blood samples, CVF, urine, and saliva. After day 9 of the cycle, daily collection of urine, CVF, and saliva continues until the end of the menstrual cycle. In accordance with one of the ovulation assessment methods, post-ovulatory monitoring is scheduled. After this estimated point, alternate-day blood sampling is performed until 10 days post-ovulation. Fourteen days post-ovulation, a visit is scheduled that includes blood sampling for beta-hCG hormone measurement and cervical brushing. At this stage, the participants in the NCG conclude their study participation.

The TCG repeats the same follow-up protocol above for up to six cycles. Throughout these cycles, clinical pregnancies are assessed via beta-hCG levels, with clinical pregnancy monitoring commencing upon detection and continuing until childbirth. Once beta-hCG has been detected, the participant moves on to the pregnancy monitoring phase. One cervical brushing is performed on day 21 post-ovulation. Following this, the participants underwent an ultrasound at 7, 11–14, 24–26, and 32–34 weeks. In addition to ultrasound monitoring, blood samples and brushings are collected during each trimester, specifically at weeks 11–14, 24, and 36. Standard prenatal care, in accordance with Chilean guidelines, is provided either by the obstetrician from the research team or by the obstetrician chosen by the participant based on their preference (Minsal, 2015). At childbirth, placental tissue, umbilical cord blood, and newborn data are collected.

### 3.3 Recruitment numbers to date

The EARLY-PREG open cohort provides a dynamic characterisation of menstrual cycle patterns and reproductive outcomes through a two-way longitudinal study. The demographic and general population characteristics of the EARLY-PREG open cohort at present are detailed in Table 1. In Table 2, we present the total number of cycles contributed by participants in the NCG and TCG.

**Table 1.**
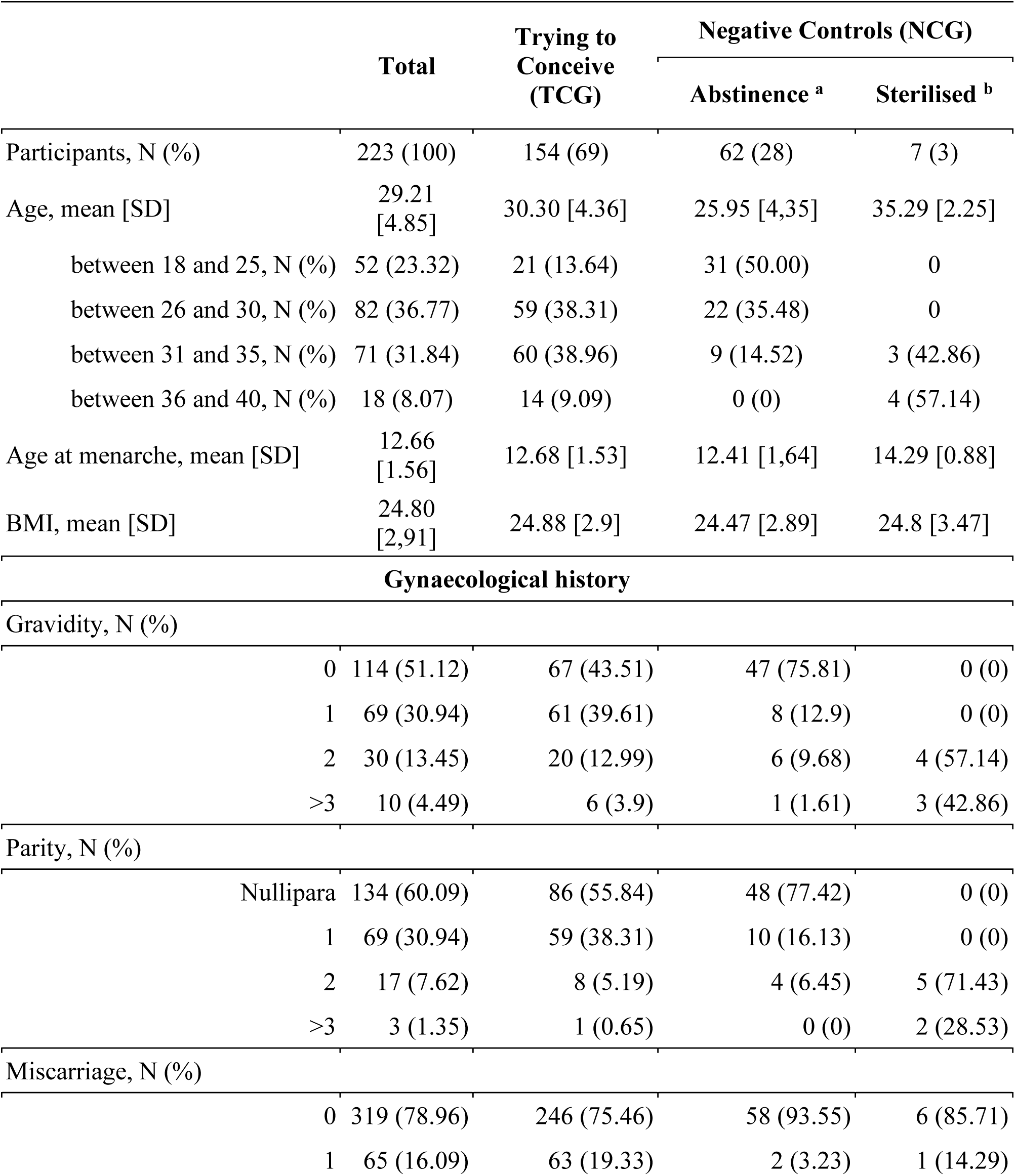
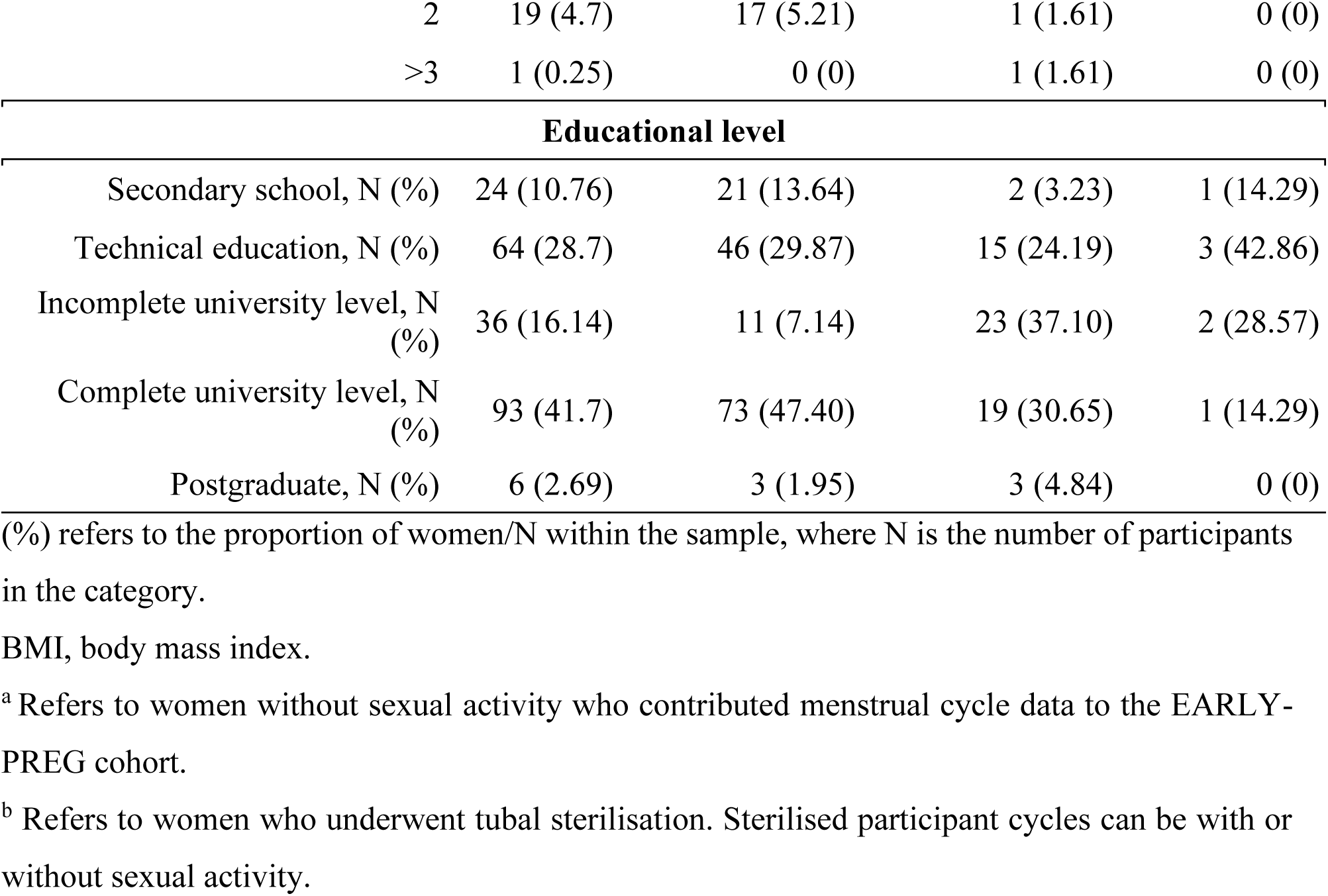
Demographic and reproductive characteristics of the enrolled participants in the EARLY-PREG cohort.

**Table 2.** Number of cycles contributed by participants per group.

|  | Volunteers |  |  | Cycles |  |
| --- | --- | --- | --- | --- | --- |
|  | Trying to Conceive (TCG) | Negative Controls (NCG) |  | Non conception | Conception |
|  |  | Abstinence | Sterilised |  |  |
| **1 cycle, N (%)** | 64 (51.61) | 53 (42.74) | 7 (5.65) | 102 | 22 |
| **2 cycles, N (%)** | 47 (82.46) | 9 (17.54) | 0 (0) | 95 | 17 |
| **3 cycles, N (%)** | 23 (100) | 0 (0) | 0 (0) | 61 | 8 |
| **4 cycles, N (%)** | 6 (100) | 0 (0) | 0 (0) | 22 | 2 |
| **5 cycles, N (%)** | 9 (100) | 0 (0) | 0 (0) | 41 | 4 |
| **6 cycles, N (%)** | 5 (100) | 0 (0) | 0 (0) | 30 | 0 |
| **Total, N** | 154 | 62 | 7 | 351 | 53 |

N represents the number of participants who contributed between 1 and 6 cycles in their respective categories.
(%) represents the proportion of contributors per group/individuals per cycle category.

### 3.4 Menstrual Cycle Description and Outcomes

We provide an overview of the participants’ menstrual cycle characteristics (Table 3). A total of 404 menstrual cycles have been documented to date, classified as either non-conception (n = 352) or conception cycles (n = 52). Among the conception cycles, 35 resulted in full-term pregnancies, whereas 17 ended in early pregnancy loss (EPL), which is defined as a miscarriage until 12 6/7 weeks (American College of Obstetricians and Gynaecologists’ Committee on Practice Bulletins-Gynaecology, 2018).

**Table 3.**
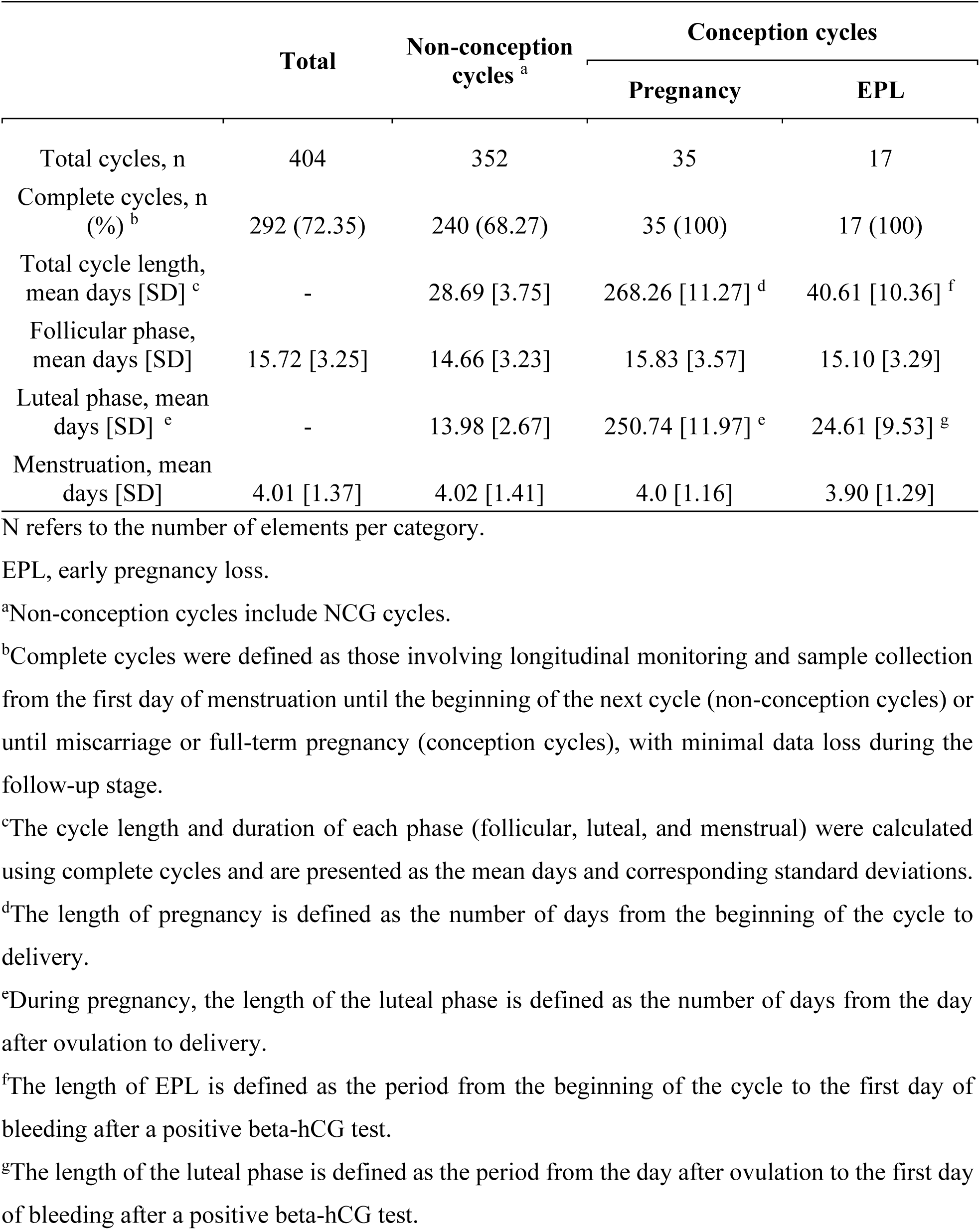
Menstrual cycle characteristics according to outcome in the EARLY-PREG cohort.

|  | Total | Non-conception<br>cycles <sup>a</sup> | Conception cycles |  |
| --- | --- | --- | --- | --- |
|  |  |  | Pregnancy | EPL |
| Total cycles, n | 404 | 352 | 35 | 17 |
| Complete cycles, n<br>(%) <sup>b</sup> | 292 (72.35) | 240 (68.27) | 35 (100) | 17 (100) |
| Total cycle length,<br>mean days [SD] <sup>c</sup> | - | 28.69 [3.75] | 268.26 [11.27] <sup>d</sup> | 40.61 [10.36] <sup>f</sup> |
| Follicular phase,<br>mean days [SD] | 15.72 [3.25] | 14.66 [3.23] | 15.83 [3.57] | 15.10 [3.29] |
| Luteal phase, mean<br>days [SD] <sup>e</sup> | - | 13.98 [2.67] | 250.74 [11.97] <sup>e</sup> | 24.61 [9.53] <sup>g</sup> |
| Menstruation, mean<br>days [SD] | 4.01 [1.37] | 4.02 [1.41] | 4.0 [1.16] | 3.90 [1.29] |

N refers to the number of elements per category.
EPL, early pregnancy loss.
<sup>a</sup>Non-conception cycles include NCG cycles.
<sup>b</sup>Complete cycles were defined as those involving longitudinal monitoring and sample collection from the first day of menstruation until the beginning of the next cycle (non-conception cycles) or until miscarriage or full-term pregnancy (conception cycles), with minimal data loss during the follow-up stage.
<sup>c</sup>The cycle length and duration of each phase (follicular, luteal, and menstrual) were calculated using complete cycles and are presented as the mean days and corresponding standard deviations.
<sup>d</sup>The length of pregnancy is defined as the number of days from the beginning of the cycle to delivery.
<sup>e</sup>During pregnancy, the length of the luteal phase is defined as the number of days from the day after ovulation to delivery.
<sup>f</sup>The length of EPL is defined as the period from the beginning of the cycle to the first day of bleeding after a positive beta-hCG test.
<sup>g</sup>The length of the luteal phase is defined as the period from the day after ovulation to the first day of bleeding after a positive beta-hCG test.

Menstrual cycles were classified based on whether ovulation took place or not as ovulatory or anovulatory, respectively. Moreover, we included a group of uncharacterised cycles in the process of classification at the time of submitting this study. Table 4 summarises the distribution of these cycle types and their corresponding outcome classifications.

**Table 4.** Quantification of menstrual cycles according to the outcomes and occurrence of ovulation in the EARLY-PREG cohort.

| Cycle type | Non-conception <sup>a</sup>, n (%) | Conception cycles |  |
| --- | --- | --- | --- |
|  |  | Pregnancy | EPL |
| **Ovulatory** <sup>b</sup> | 95 | 35 | 17 |
| **Anovulatory** | 15 | - | - |
| **Uncharacterised** <sup>c</sup> | 131 | - | - |

EPL, early pregnancy loss.
<sup>a</sup>Non-conception cycles include NCG cycles.
<sup>b</sup>Ovulation was assessed by ecography and LH measurement (fertility monitor or test strips and surge calculation; Godbert et al., 2015).
<sup>c</sup>Samples without LH measurement or ultrasound, and ovulation confirmation based on progesterone measurement

### 3.5 Structured Longitudinal Biobank and Performance

A total of 405 menstrual cycles have been monitored in the EARLY-PREG cohort biorepository so far, which includes 6,406 CVF samples, 5,120 urine samples, 1,467 saliva samples, 928 blood samples, and 293 brushing samples collected throughout the menstrual cycles. Additionally, 17 placenta and umbilical cord blood samples have been obtained from term pregnancies, providing a valuable resource for studying perinatal outcomes. 292 complete menstrual cycles were biologically sampled from the monitored cycles: 43 from the NCG and 249 from the TCG. The NCG samples collected from the EARLY-PREG cohort included 208 blood samples, 832 CVF samples, 876 urine samples, 279 saliva samples, and 26 cervical brushing samples. The cohort cycles have been further classified into counterfactual and non-counterfactual groups to facilitate reproductive studies on pregnancy, early pregnancy loss, and non-conception. Table 5 presents the total number of samples collected from the complete TCG cycles (excluding those from NCG). Within the TCG, 78 cycles were classified as counterfactual, and 171 were classified as non-counterfactual.

**Table 5.**
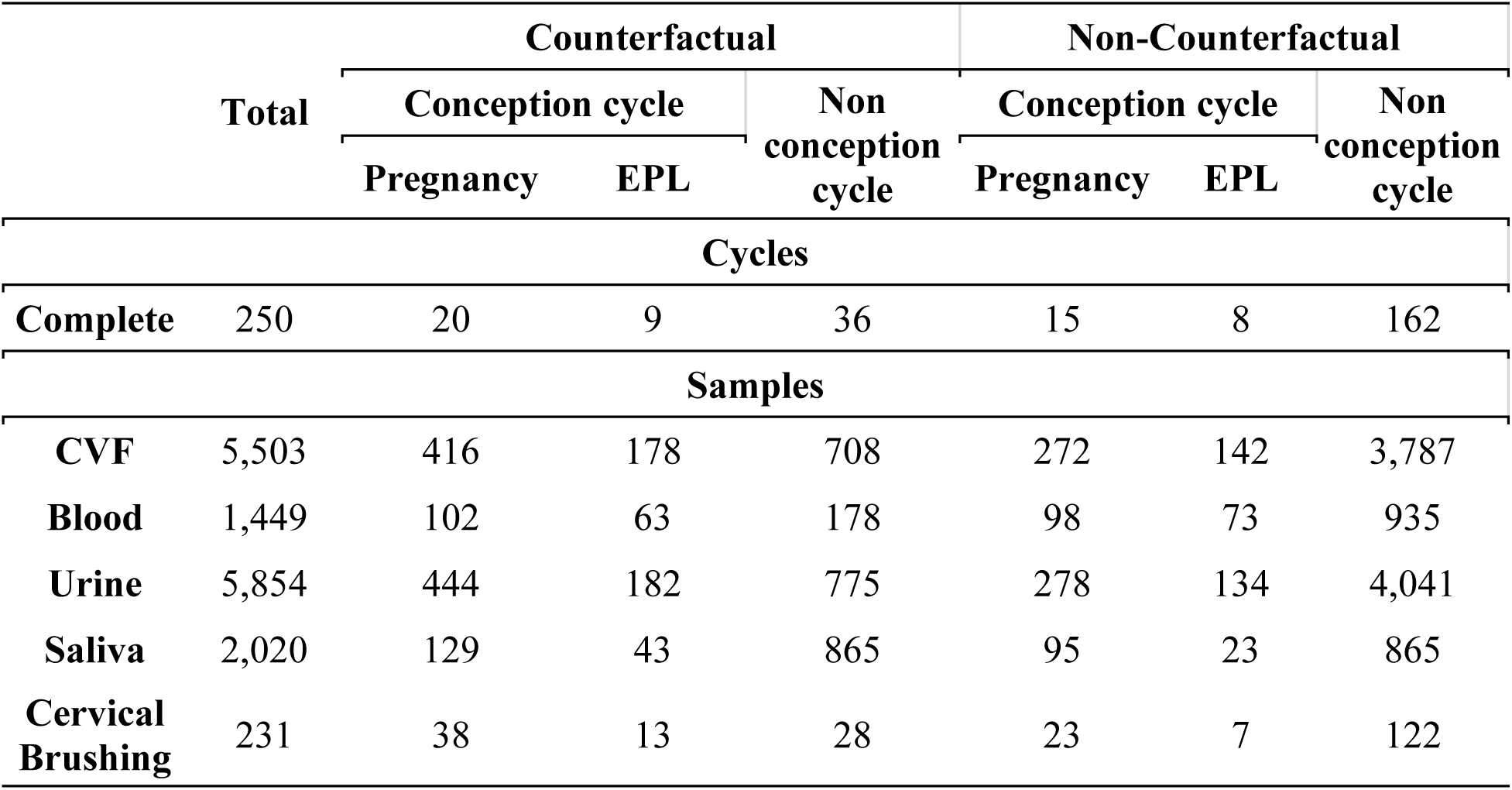
Classification of complete cycles of the TCG in the EARLY-PREG cohort. EPL, early pregnancy loss; CVF, cervicovaginal fluid.

### 3.6 Biospecimen Collection by Cycle Day and Type

The collection of biospecimens has been categorised by cycle day and type so far, as shown in Figure 5. The timeframes enabled the evaluation of samples of non-conception and conception cycles (pregnancy and early pregnancy loss). We highlight the presence of samples collected during the first two weeks after conception in this longitudinal cohort study.

**Figure 5.**
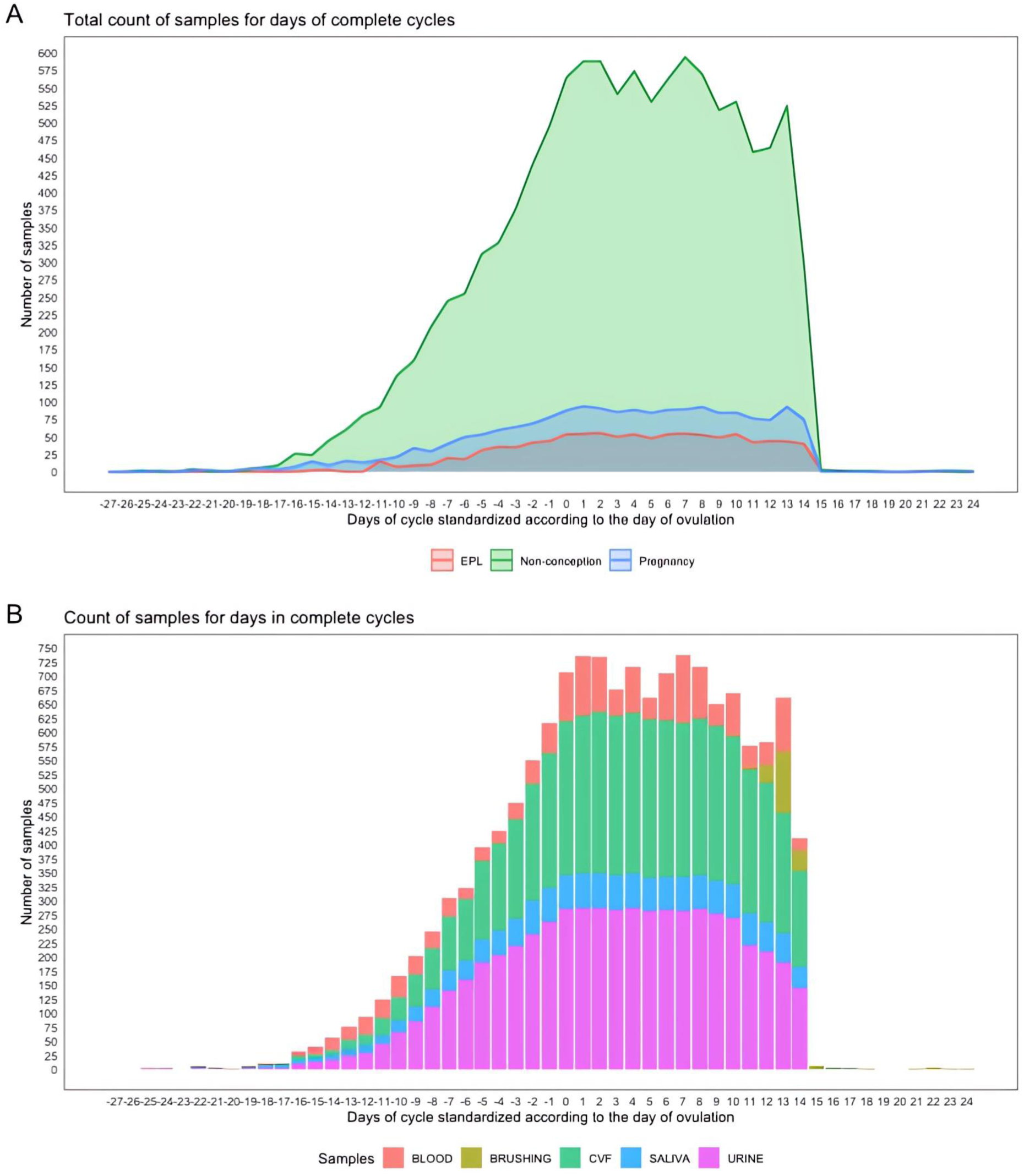
Total sample count per day across cycles in complete cycles. The x-axis represents the days relative to the cycle, standardised according to the day of ovulation, whereas the y-axis indicates the total number of collected samples. A. Total sample count for EPL, pregnancy, and non-conception cycles. B. Counts of samples per day in total cycles. Sample types are colour-coded: CVF (green), blood (red), urine (magenta) saliva (light blue), and brushing (lime green). EPL, early pregnancy loss; CVF, cervicovaginal fluid.

### 3.7 Biospecimen Collection Performance

We have evaluated so far the performance of the structured longitudinal biobank by comparing the number of expected samples to the actual samples obtained during the specified windows of interest. The efficiency of sample collection is determined by calculating the percentage of completeness within the windows via Formula (1), and we adjusted the CVF and urine according to Formula (2).

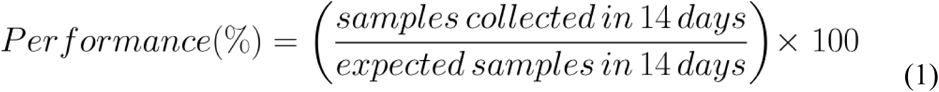

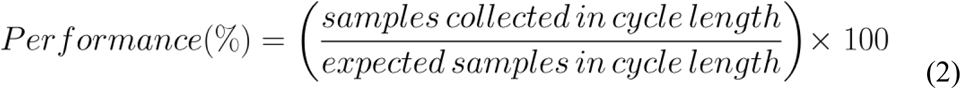

Overall compliance with sample collection during defined windows of interest has been high for most biospecimens. For CVF, 3,956 out of 4,380 expected samples were collected (90.3%) in the window from day 0 to 14, and 96.15% were collected in the window adjusted for each patient’s luteal phase length; for urine samples, 3,880 out of 4,380 samples were collected (88.6%) during the period from days 0 to 14 across study cycles, and 94.29% were collected in the window adjusted for each patient’s luteal phase length. In contrast, compliance was lower for saliva (collected in wave three and, in some cases, in wave two), with 89.22% in the window adjusted for each patient’s luteal phase length (2). For blood, 1,499 of the 3,600 expected samples were collected (41.6%) across seven scheduled collections from day 0 to 10 of the window.

Throughout the study, a total of 287 out of 462 brushing samples were collected from TCG and NCG. Brushing was expected at least once per participant, depending on the study aim and reproductive outcome. During the pregnancy monitoring stage of TCG, 72 out of 105 expected brushings were collected, resulting in a compliance rate of 68.6%. Among conceptions and EPL, brushing was expected once or twice (±14 and ±21 days post-ovulation), depending on gestational progression. Of the 347 expected samples, 215 were obtained, yielding a compliance of 62.0%.

## 4. Strengths and limitations

- To our knowledge, the EARLY-PREG cohort is the first study design aiming to research embryo‒mother molecular exchange with a preconception, longitudinal, bidirectional, and counterfactual approach. This unique strategy allows each participant to serve as their own control to compare local and systemic effects of the presence or absence of a naturally conceived embryo, minimising interindividual variability during ultra-early pregnancy.
- A key strength of this study is its prospective open design, which allows follow-up across participants for one or more menstrual cycles, providing detailed data across critical reproductive phases, especially the first two weeks after conception.
- We established a comprehensive multisample biobank including CVF, urine, blood, saliva and cervical brushings samples, as well as placenta and umbilical cord blood to support future research in human reproduction.
- In the EARLY-PREG cohort, biological samples were collected exclusively from women. Like other preconception and pregnancy cohorts, our data on male partners are limited to information from participating women or a short baseline survey.
- Another limitation is the lack of prior clinical records for the participants. During the recruitment and baseline surveys, the participants self-reported their medical conditions and family medical history; however, this information was not verified against existing medical records.
- Fertilisation events may have taken place without successful implantation, with beta-hCG testing not detecting these cases. Advances in proteomic profiling in CVF from our biorepository could allow the identification of markers that recognise these events.
- Like many voluntary longitudinal studies, the cohort is susceptible to selective recruitment, which, combined with geographical constraints, may limit the representativeness of the findings for broader populations.

## 5. Future plans

The EARLY-PREG cohort biorepository has reached a sufficient size and number of biological samples to begin characterising the longitudinal proteomes of the conception and non-conception cycles. The extensive collection of women’s and maternal biospecimens opens the possibility for future advances in the study of longitudinal changes in the proteome throughout key stages of female human reproduction. Additionally, biological samples have been collected from full-term pregnancies, providing a valuable resource for further research on perinatal outcomes.

We project to identify local and systemic maternal responses associated with the embryo‒mother molecular dialogue in the early stages of pregnancy. To this end, we will analyse biological samples from the EARLY-PREG cohort using a proteomics approach, following the proposed design and workflow of Figure 6, in which consecutive days during the early pregnancy stage are compared between participants who achieved pregnancy and their respective previous non-conception counterfactual cycles.

**Figure 6.**
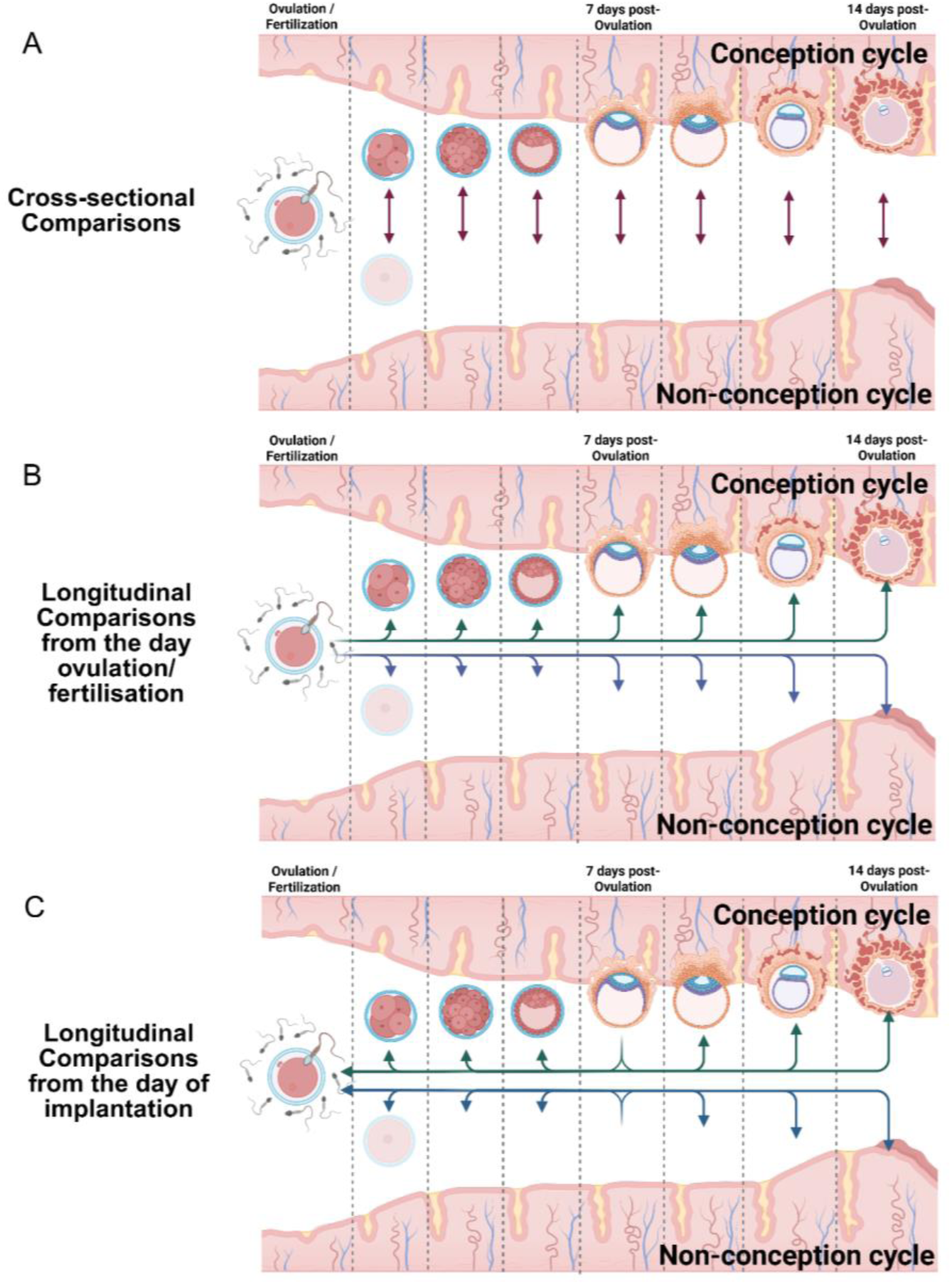
Proposed designs for analytic comparisons across the cycles and samples of the EARLY-PREG cohort. Panel A illustrates a cross-sectional comparison approach, where each day is compared with its equivalent in both conception and non-conception cycles, following synchronisation (based on LH surge or beta-hCG). Panels B and C present longitudinal comparison approaches. In panel B, days are aligned according to the day of ovulation/fertilisation, synchronised using the LH surge, in both conception and non-conception cycles. Panel C uses the same approach but aligns days based on implantation. This method employs beta-hCG as an implantation marker, which serves as the reference point for synchronisation under this standard. All comparisons presented are based on a counterfactual model of the EARLY-PREG open cohort.

In a companion study (Hernandez *et al.,* 2025), we validated a first data-independent acquisition parallel accumulation–serial fragmentation (dia-PASEF) method for CVF MS-based proteomics analysis in this biofluid. Based on this work, we will characterise the molecular communication between the embryo and the mother. This will be complemented by subsequently analysing other biospecimens such as blood and urine. Our study is intended to cover the period from ovulation to post-embryo implantation (day 0 to 14). The primary objective of this stage is to identify the proteome signature of conception by comparing the CVF proteome of participants who successfully conceived with their respective counterfactual non-conception cycles, while also quantifying associated biological processes and potential biomarkers. The biospecimens collected will also enable us to characterise additional key periods of human reproduction, including the fertile window (−5 to 0 days) (Wilcox *et al*., 1995), the periovulatory window (−3 to 3 days), and the implantation window (7 to 10 days), synchronised according to the day of ovulation or embryo implantation (see figure 4).

Finally, a new recruitment wave of the EARLY-PREG study is set to begin exploring maternal immunoregulation and the microbiome during the ultra-early stages of pregnancy. This new wave will involve a multi-omic approach, incorporating advanced techniques such as flow cytometry, proteomic, metaproteomic, and metagenomic analyses. We will collect cervical fluid, endometrial biopsy, and stool samples, in addition to baseline samples, including CVF, urine, blood, saliva, and cervical brushing. This design facilitates comparisons following the proposed counterfactual framework. The results obtained may contribute to future research on maternal health and fertility and represent an important step towards future studies of biomarkers of ultra-early pregnancy.

## 6. Collaboration

The datasets generated and/or analysed during the current study are not publicly available because of the sensitivity of the participant material. Data requests can be sent to the corresponding author.

## 7. Contributors

E.S.K. conceived the original study using a two-way open longitudinal design. C.V. and J.S. contributed to the implementation of all three recruitment waves. M.H., E.C. and B.A. contributed to the implementation and maintenance of the biobank. V.L., G.N., and C.V. contributed with database construction and data analysis. D.A., V.L., G.N., and B.A. drafted the working manuscript, and E.S.K., C.V., I.F-M., and G.N. reviewed and commented on drafts. All the authors approved the final manuscript. E.S.K. is the guarantor of the study, accepts full responsibility for the research, has full access to the data, and controls the decision to publish.

## 9. Acknowledgments

We wish to express our deepest gratitude to all the participants and couples who took part in the complex and demanding EARLY-PREG open cohort. We would also like to thank all midwives, obstetricians, nurses, other medical staff, laboratory and research personnel, and administrative staff for the management of participants and their pivotal role in data collection. We thank Estefania Nova-Lamperti for her support in figure design, and Ivo Fierro-Monti for advice and revision of language.

## 11. Data availability statement

The data are available upon reasonable request. All data relevant to the study are included in the manuscript.

